# Development of the TBQ+D: A Novel Patient-Reported Measure of The Burden of Digital Care

**DOI:** 10.1101/2025.05.12.25327446

**Authors:** Misk Al Zahidy, Kerly Guevara, Sue Simha, Mariana Borras-Osorio, Megan E. Branda, Viet-Thi Tran, Jennifer L. Ridgeway, Victor M. Montori

## Abstract

**Background:** Patients with diabetes often manage complex treatment regimens that increasingly include the use of digital medicine tools. While several instruments measure treatment burden, none capture the specific burden introduced by digital medicine tools used in self-care. Development of patient reported measures to capture digital treatment burden are needed to assess the lived experiences and challenges patients face when using digital medicine tools.

**Objective:** To engage patients and clinical experts in adapting an existing measure of treatment burden—the Treatment Burden Questionnaire (TBQ). The adapted instrument underwent cognitive testing and refinements to ensure it could capture the burden of using digital medicine tools in the self-management and care of diabetes.

**Methods:** This two-phase study was conducted with adults with diabetes and other chronic conditions at the Division of Endocrinology at Mayo Clinic in Rochester, Minnesota. First, we mapped themes from prior concept elicitation interviews to existing TBQ items to identify content gaps related to digital burden. Based on these gaps, the study team and expert panel followed an item development guide to generate new items and adapt existing ones to better reflect the workload and burdens imposed by digital medicine tools. The resulting adapted instrument underwent three rounds of cognitive testing with adult patients living with diabetes, using a think-aloud protocol to assess clarity, relevance, and comprehensiveness. Results of cognitive testing informed iterative refinements across three rounds of interviews, leading to improved clarity, reduced redundancy, and improved relevance of items.

**Results:** The final TBQ+D retained the structure of the original 15-item TBQ and included 8 new items and modified 8 extant ones to capture burden of digital care (e.g., syncing issues, discomfort from sensors, and device malfunctions). Cognitive testing demonstrated strong content relevance and patient comprehension.

**Conclusion:** The TBQ+D appears able to measure digital treatment burden in patients with diabetes. Planned next steps include field testing the instrument to test validity hypotheses, and if successful, extending this evaluation to diverse populations and clinical and research settings.

## BACKGROUND

Chronic conditions, such as diabetes, affect over 100 million adults in the United States^1^, placing a significant workload on patients to access care, follow treatment plans, and engage in daily self-management.^2–6^ For patients living with diabetes, this often includes monitoring blood glucose, making self-management decisions, implementing insulin dosing, and managing food choices and exercise.^7–10^ This cumulative workload, and its impact on a patient’s quality of life is known as treatment burden.^7,11–14^ Higher treatment burden is linked with poorer health outcomes, including reduced adherence to care plans and increased hospitalization rates.^7,15^

In response to these challenges, digital medicine tools and applications that aim at supporting self-management—such as continuous glucose monitors (CGMs), insulin pumps, patient portals, mobile apps, and telemedicine—have been increasingly integrated into diabetes care.^16–20^ These offer potential benefits by automatic or supporting self-management tasks.^17,19–23^ However, they can also introduce new work and source of strain, including technical malfunction, usability issues, and information overload, particularly those less familiar with or with limited access to digital technology^24,25^. In our prior work patients describe how digital tools sometimes complicated rather than eased the workload of managing chronic illness.^26^

Despite this growing digital landscape, existing instruments that assess treatment burden such as the Treatment Burden Questionnaire (TBQ) and Patient Experience with Treatment and Self-management (PETS), do not capture the burden associated with digital tools. ^5,13,15,24,27–29^ This gap hinders our ability to understand and address the challenges patients face by using digital medicine tools.^26^

To fill this gap and capture the burden of using digital medicine tools, the Treatment Burden Questionnaire Plus Digital (TBQ+D) was developed to capture the burden of treatment in patients living with diabetes, including the burden of digital medicine tools.

## METHODS

The study builds on previously reported concept elicitation interviews of patient work and experience of using digital tools.^26^ This report includes two phases: (1) mapping concept elicitation themes to the existing TBQ items and generating new or modified items, and (2) cognitive testing and refinement to address gaps.

### Participants

For both of the phases, adult patients aged 18 years or older with a diagnosis of type 1 or type 2 diabetes and attending the diabetes clinic in the Division of Endocrinology at Mayo Clinic (Rochester, Minnesota) were eligible to participate if they used at least one digital medicine tool for diabetes management (e.g. glucometer, insulin smartpens, pumps, etc.), were proficient in English, and were able to complete informed consent (i.e., had no major functional impairment). Patients whose caregivers were the primary users of digital tools or who had significant cognitive or sensory impairments were excluded.

Eligible patients were identified from the daily appointment calendar and approached in-person by study staff. Recruitment aimed to maximize diversity in diabetes type (type 1 or type 2) and intensity of digital tool use which was measured using the Digital Medicine Tools Intensity Scale—a 7-point scale ranging from “none” to “maximal” (**Figure 2**).^26^

**Figure 1:**
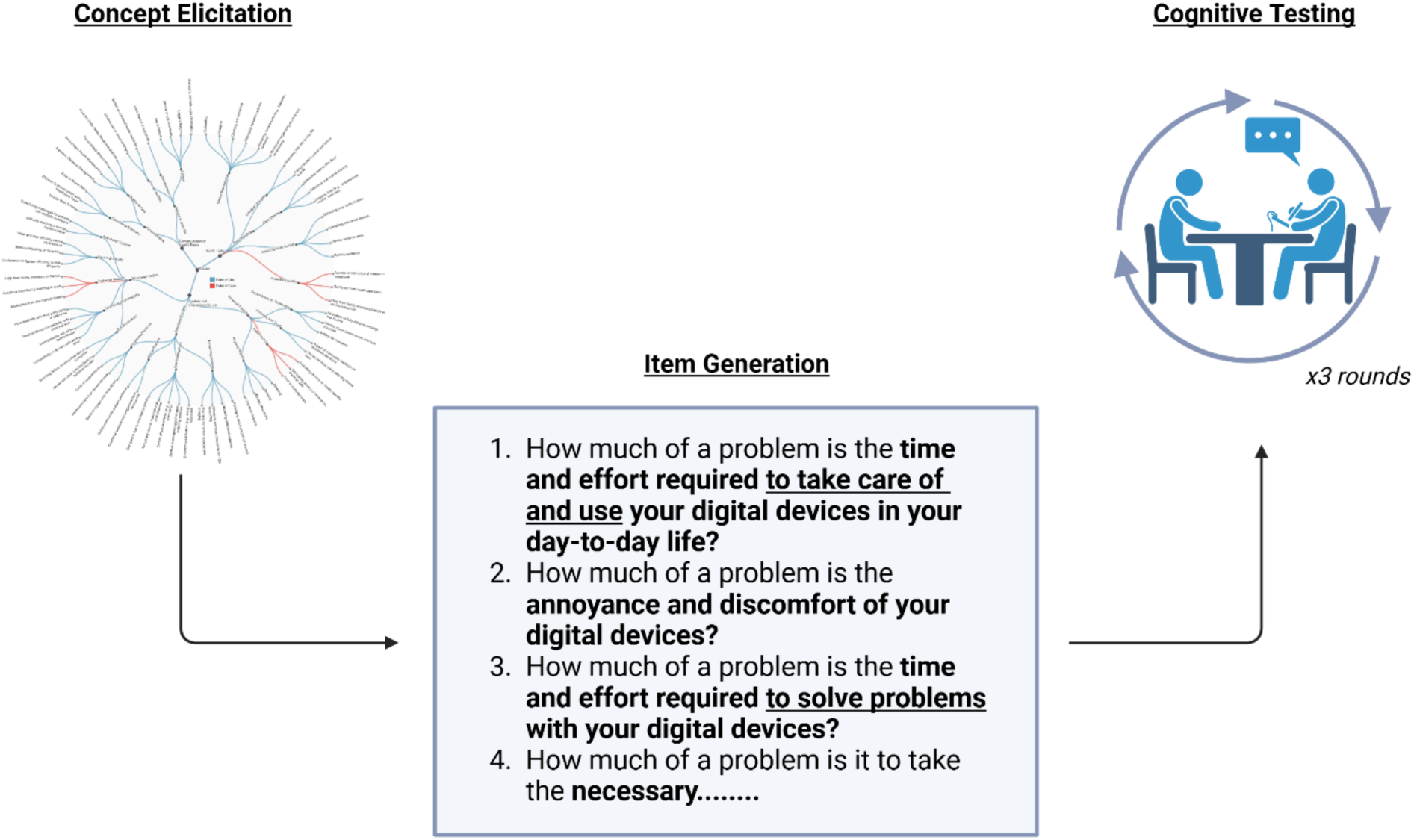
The development process.

**Figure 2:**
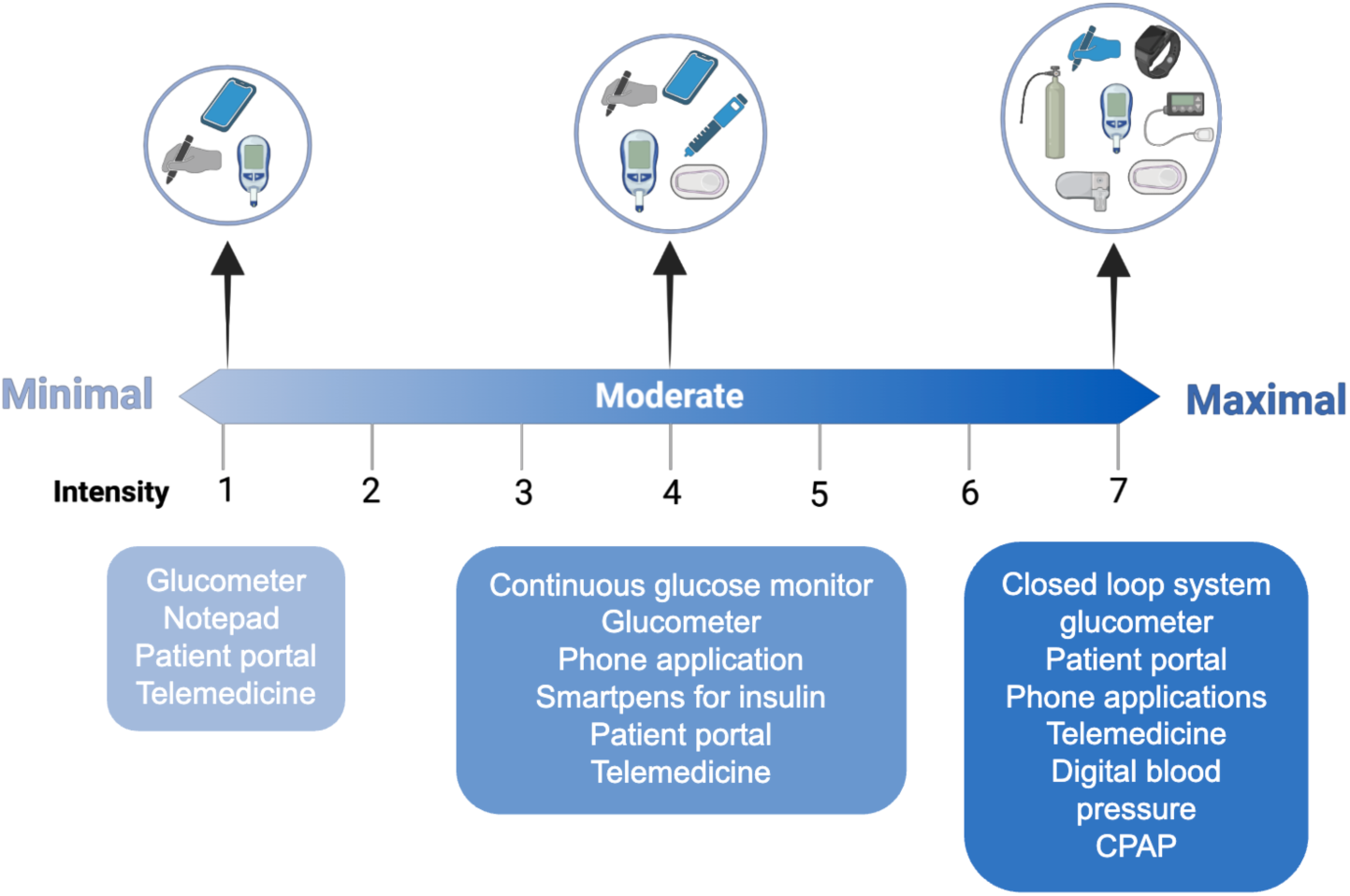
Medicine Tools Intensity Scale.

### Item Generation

To guide item generation, we conducted a structured mapping exercise linking qualitative data from the concept elicitation study^26^ to items in the original TBQ **(Table 1).** Using content analysis, two team members reviewed patient responses to each TBQ item and coded themes that reflected how digital tools impacted the same burden domains. When novel burden domains emerged that were not addressed by existing TBQ items, new items were generated guided by the original item stems and adapted to reflect the digital context. A multidisciplinary expert panel reviewed all proposed additions and modifications to ensure conceptual clarity and relevance prior to cognitive testing.

**Table 1:**
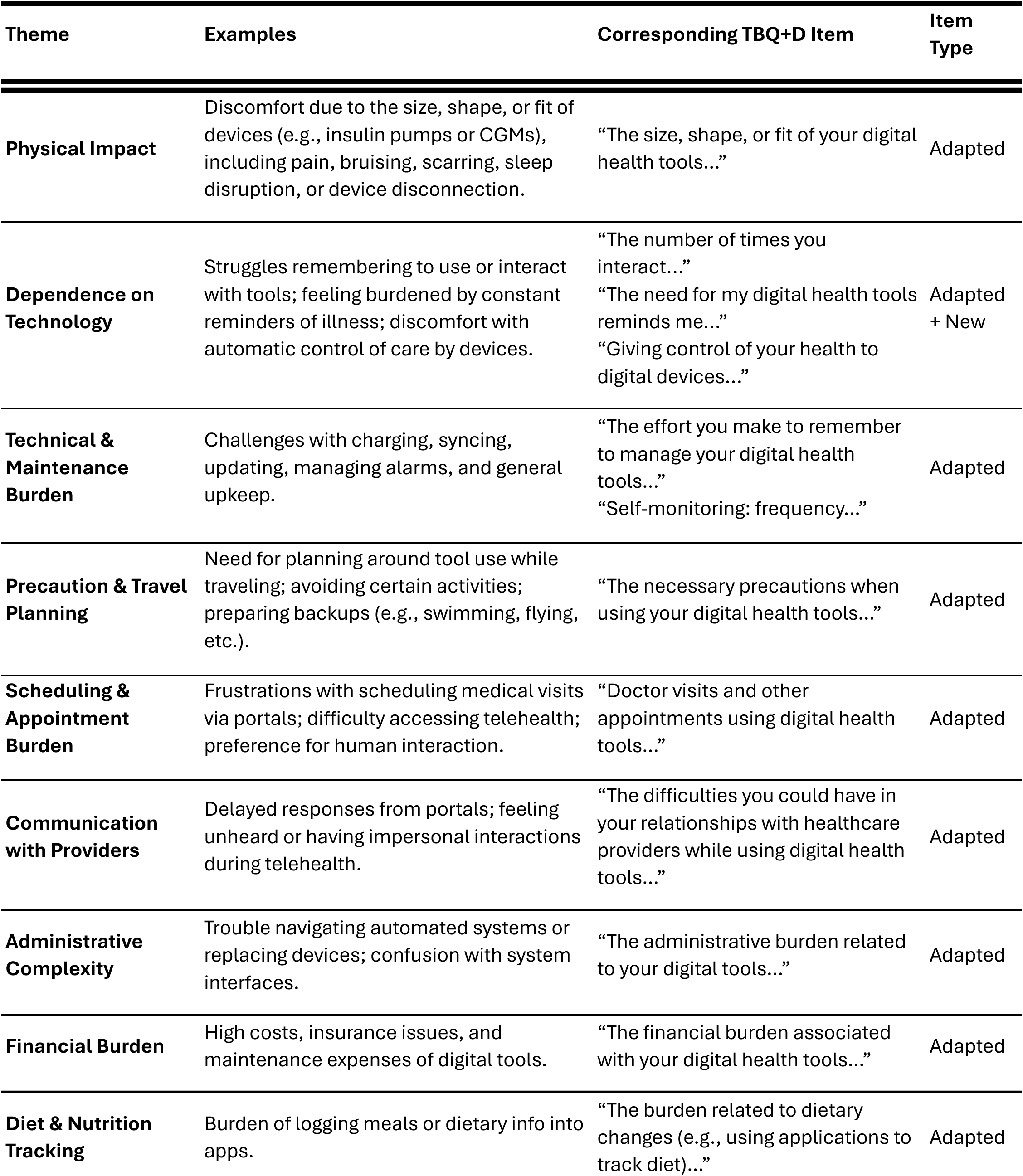
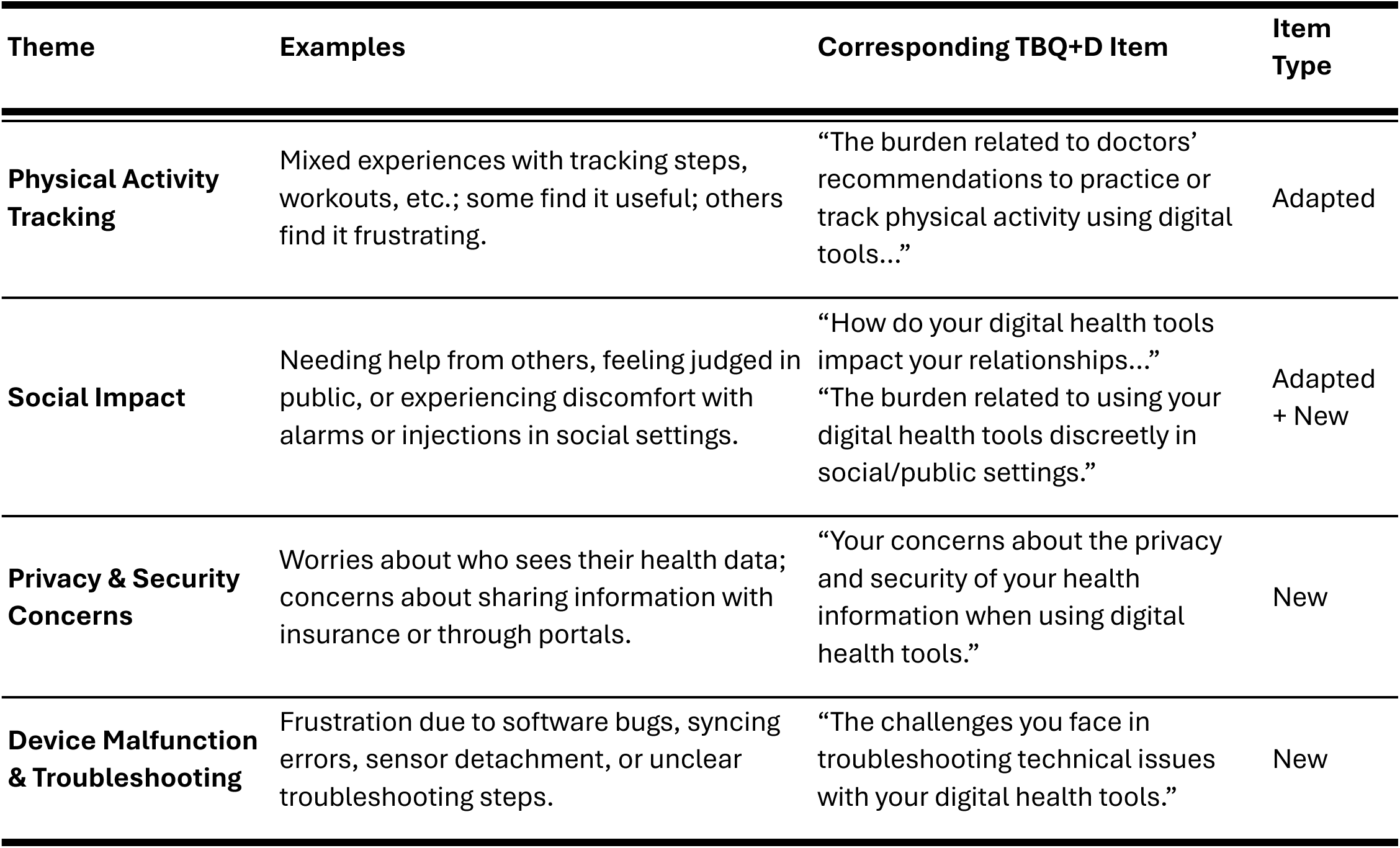
Mapping burden of digital tool themes to TBQ+D items.

### Cognitive Testing

Once the initial draft of the TBQ+D was complete, a new sample of patients were recruited to pre-test the TBQ+D over three rounds of cognitive interviews, each involving up to 10 participants. Participants completed the TBQ+D during in-person private sessions using a think-aloud protocol by the same trained researcher (M.A.Z.). As they responded to each item, participants verbalized their thought processes, interpretations, and any confusion they experienced. This approach allowed researchers to assess the clarity, relevance, and wording of the questionnaire items. Participants were asked about difficulties in understanding any items, suggestions for improving item wording or content, and whether any aspects of their digital tool use experience were not captured by the questionnaire.

### Data Handling

Survey responses, demographic, and clinical data were managed using REDCap electronic data capture tools hosted at Mayo Clinic (UL1TR002377).

### Data Analysis

As previously published, themes from the concept elicitation phase were derived from patient interviews exploring the work and experience of using digital medicine tools.^26^ In the current study, two members of the research team reviewed these published themes and mapped them to items from the TBQ to evaluate item coverage and identify content gaps. When burden domains related to digital care—such as syncing errors, device alarms, technical malfunctions, or data management—were not adequately addressed by existing TBQ items, new items were generated using patient language and contextual detail.

For the cognitive testing phase, data from the think-aloud interviews were analyzed for patient interpretations of specific questions and overall survey clarity, applicability, and completeness. Notes and transcripts were reviewed after each round to identify issues with item interpretation, clarity, or relevance. Revisions were made iteratively following each round, and refinement continued until no new changes were suggested, indicating saturation.

### Ethical consideration

The Mayo Clinic Institutional Review Board (IRB Number 23-007631) approved all procedures. Informed consent was obtained from each participant. Written consent was captured prior to interviews.

## RESULTS

### Item Generation

Themes informing item development were derived from a previously published concept elicitation study.^26^ In that work, patients described the effort, complexity, and consequences of using digital tools for self-management, identifying sources of burden such as syncing failures, technical malfunctions, device-related discomfort, and frequent alerts.

For example, the TBQ item on “pain or discomfort from device” was expanded to include discomfort specific to sensor insertions, allergic reactions to adhesives, and scarring from continuous glucose monitors. Other examples, such as device malfunctions, syncing errors, and disruptions from frequent alarms, led to the generation of new items.

Informed by these themes, eight new items were developed, and several original TBQ items were refined to incorporate digital-specific examples (e.g., discomfort from sensors or issues with online scheduling). **Table 1** presents a theme-to-item mapping that illustrates how patient feedback guided the adaptation process.

### Cognitive Testing

Between April and May 2024, 24 eligible patients were approached to participate in cognitive testing, with 20 consenting (median age: 57 years, IQR: 36–67; 60% female; 40% with type 2 diabetes) **(Table 2).** Four patients declined due to time constraints.

**Table 2:**
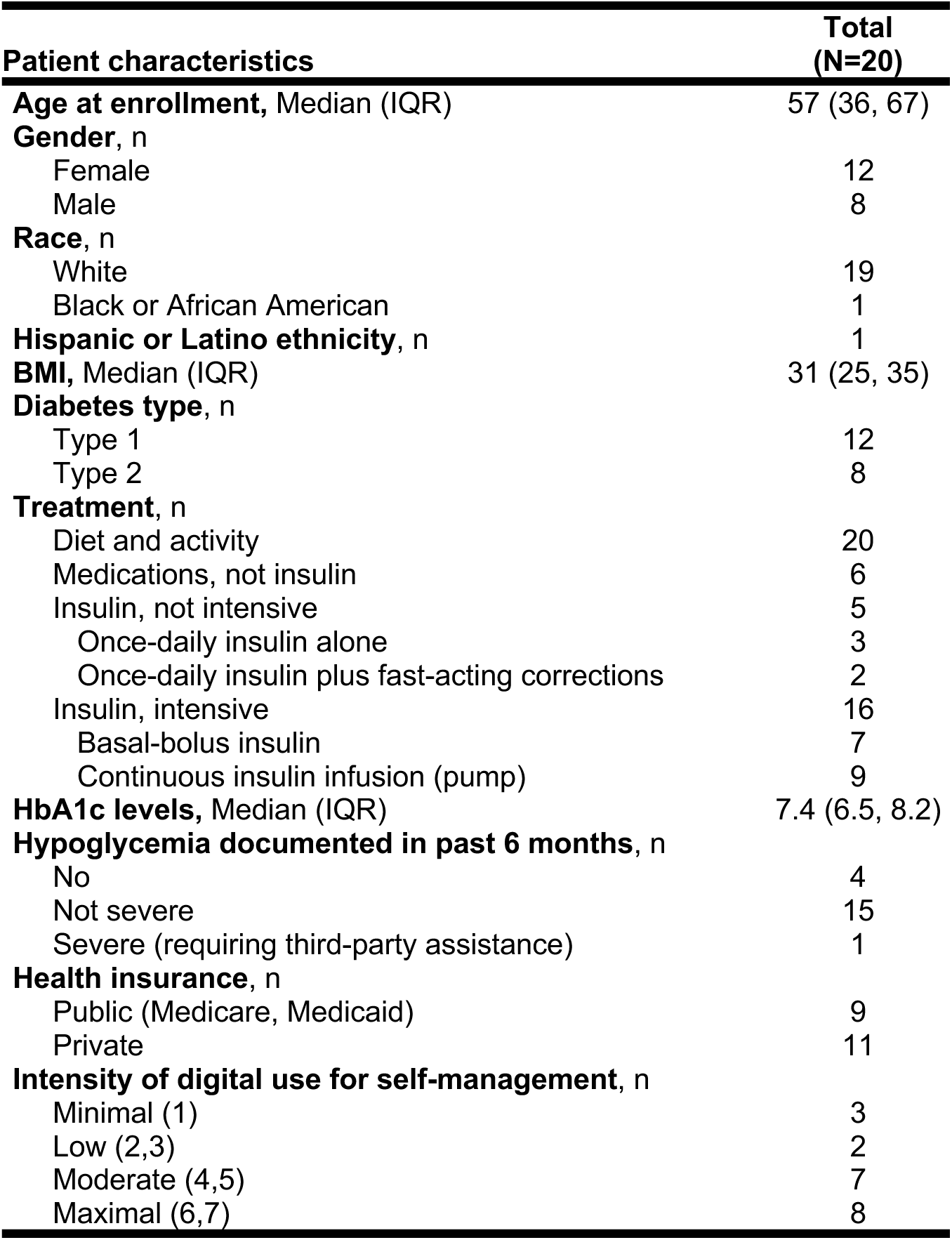
Patient characteristics.

Participants represented a range of digital medicine tool use intensities, with 8 (40%) categorized as maximal-intensity users.

The TBQ+D was refined across three rounds of cognitive interviews with up to 10 participants per round. Revisions after each round are summarized in **Table 3**.

**Table 3:** Summary of item revisions across cognitive testing rounds.

| Round | Key Feedback | Resulting Changes |
| --- | --- | --- |
| **Round 1** | Confusion with multi-faceted items (e.g., combining pills and injections into a single item). | Split items into distinct, focused items. |
|  | Misinterpretation of phrasing in original TBQ items (e.g., frequency misunderstood as the primary issue). | Moved the stem “how much of a problem” into each item to clarify intent. |
| **Round 2** | Overlap between original TBQ items and digital module questions (e.g., discomfort with wear, interruptions). | Adjusted items to remove redundancy and ensure digital module examples addressed device-specific burdens. |
|  | Confusion caused by combining "frequency" with "finding healthcare providers." | Split questions to separately address frequency/time and healthcare access (e.g., doctor visits). |
|  | Suggestions to include more examples of digital devices (e.g., smart nebulizer, at-home dialysis). | Added these examples to relevant questions for completeness. |
|  | “Bluetooth” preferred over “syncing” for describing device-related tasks. | Revised examples to reflect patient-preferred terminology. |
| **Round 3** | No new feedback indicating major issues. | Finalized item list for field testing. |

Refinements focused on improving clarity, resolving redundancy, and enhancing item relevance. Revisions were completed when no additional changes were suggested, indicating saturation. **Table 4** presents the final version of the TBQ+D.

**Table 4:**
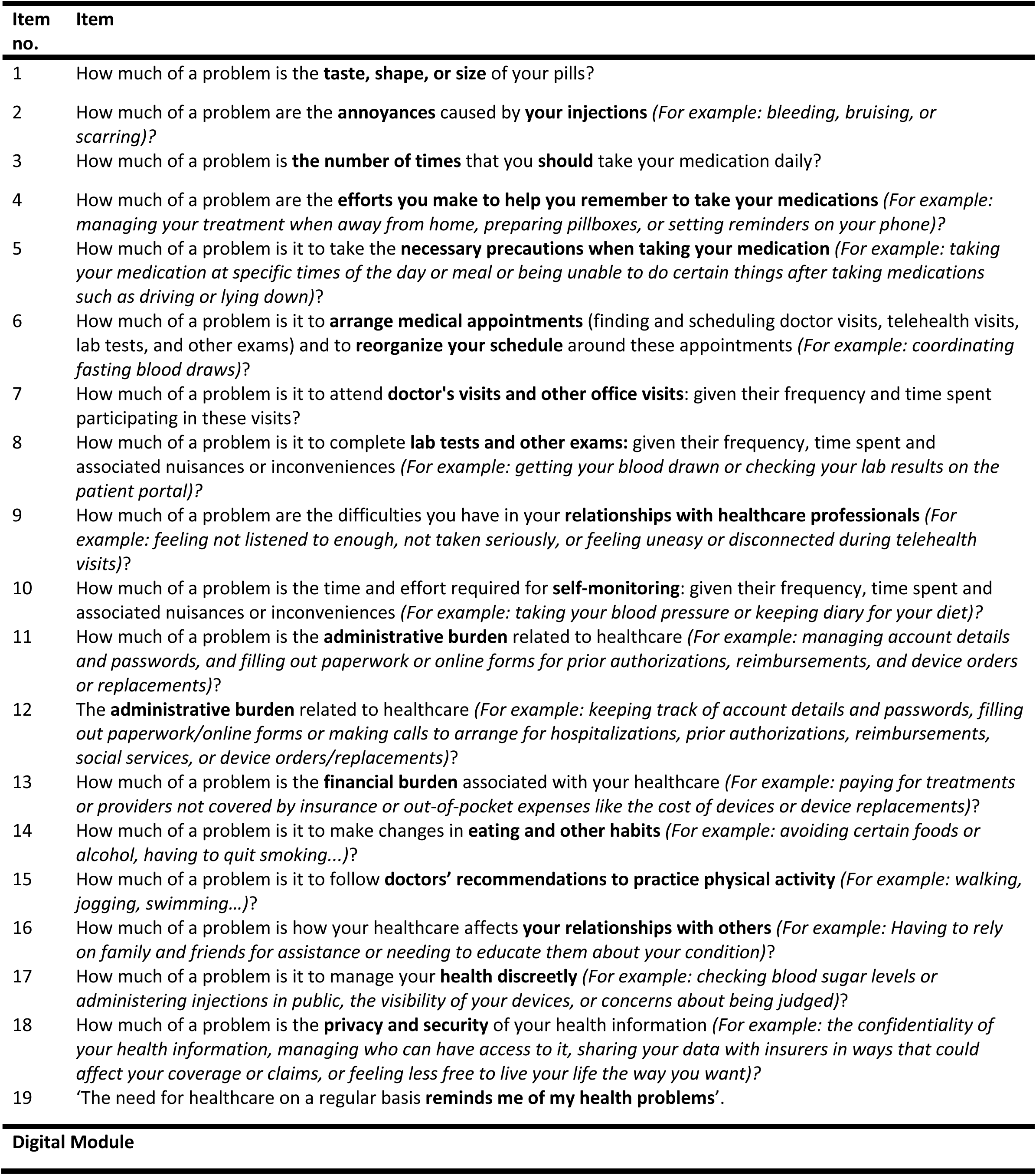
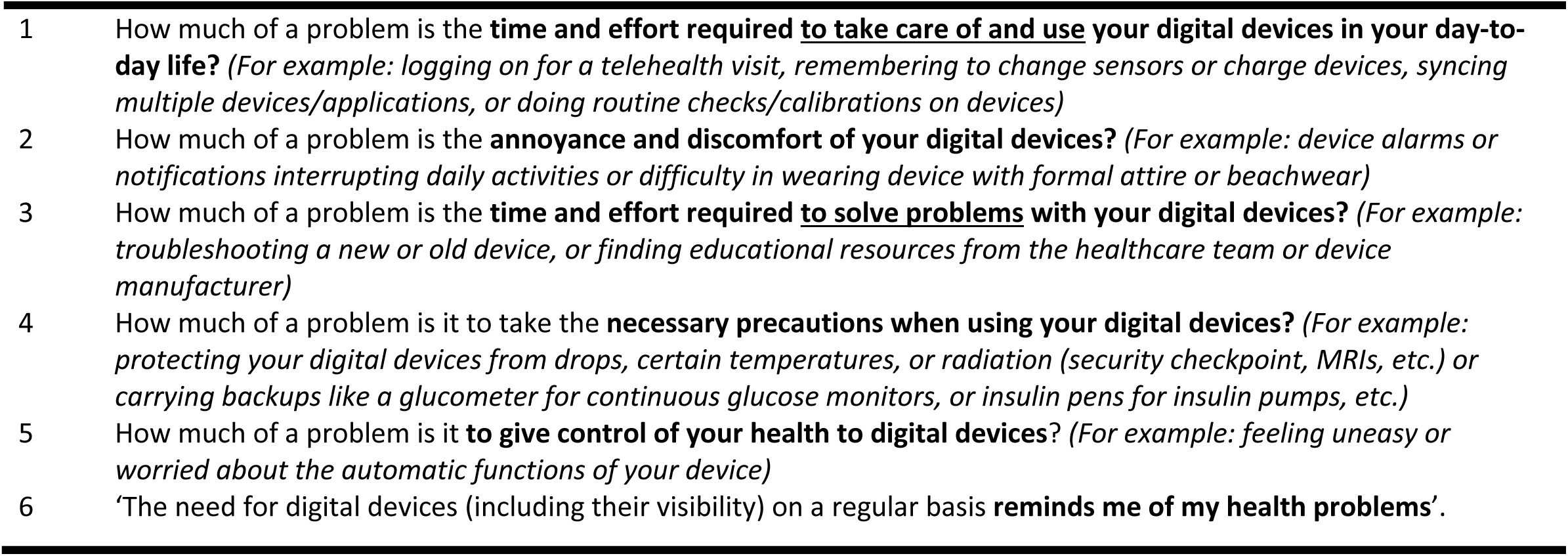
Finalized items of the TBQ+D.

| Item no. | Item |
| --- | --- |
| 1 | How much of a problem is the **taste, shape, or size** of your pills? |
| 2 | How much of a problem are the **annoyances** caused by **your injections** *(For example: bleeding, bruising, or scarring)?* |
| 3 | How much of a problem is **the number of times** that you **should** take your medication daily? |
| 4 | How much of a problem are the **efforts you make to help you remember to take your medications** *(For example: managing your treatment when away from home, preparing pillboxes, or setting reminders on your phone)?* |
| 5 | How much of a problem is it to take the **necessary precautions when taking your medication** *(For example: taking your medication at specific times of the day or meal or being unable to do certain things after taking medications such as driving or lying down)?* |
| 6 | How much of a problem is it to **arrange medical appointments** (finding and scheduling doctor visits, telehealth visits, lab tests, and other exams) and to **reorganize your schedule** around these appointments *(For example: coordinating fasting blood draws)?* |
| 7 | How much of a problem is it to attend **doctor's visits and other office visits**: given their frequency and time spent participating in these visits? |
| 8 | How much of a problem is it to complete **lab tests and other exams**: given their frequency, time spent and associated nuisances or inconveniences *(For example: getting your blood drawn or checking your lab results on the patient portal)?* |
| 9 | How much of a problem are the difficulties you have in your **relationships with healthcare professionals** *(For example: feeling not listened to enough, not taken seriously, or feeling uneasy or disconnected during telehealth visits)?* |
| 10 | How much of a problem is the time and effort required for **self-monitoring**: given their frequency, time spent and associated nuisances or inconveniences *(For example: taking your blood pressure or keeping diary for your diet)?* |
| 11 | How much of a problem is the **administrative burden** related to healthcare *(For example: managing account details and passwords, and filling out paperwork or online forms for prior authorizations, reimbursements, and device orders or replacements)?* |
| 12 | The **administrative burden** related to healthcare *(For example: keeping track of account details and passwords, filling out paperwork/online forms or making calls to arrange for hospitalizations, prior authorizations, reimbursements, social services, or device orders/replacements)?* |
| 13 | How much of a problem is the **financial burden** associated with your healthcare *(For example: paying for treatments or providers not covered by insurance or out-of-pocket expenses like the cost of devices or device replacements)?* |
| 14 | How much of a problem is it to make changes in **eating and other habits** *(For example: avoiding certain foods or alcohol, having to quit smoking...)?* |
| 15 | How much of a problem is it to follow **doctors' recommendations to practice physical activity** *(For example: walking, jogging, swimming...)?* |
| 16 | How much of a problem is how your healthcare affects **your relationships with others** *(For example: Having to rely on family and friends for assistance or needing to educate them about your condition)?* |
| 17 | How much of a problem is it to manage your **health discreetly** *(For example: checking blood sugar levels or administering injections in public, the visibility of your devices, or concerns about being judged)?* |
| 18 | How much of a problem is the **privacy and security** of your health information *(For example: the confidentiality of your health information, managing who can have access to it, sharing your data with insurers in ways that could affect your coverage or claims, or feeling less free to live your life the way you want)?* |
| 19 | 'The need for healthcare on a regular basis **reminds me of my health problems**'. |
| Digital Module |  |
| 1 | How much of a problem is the **time and effort required to take care of and use your digital devices in your day-to-day life?** *(For example: logging on for a telehealth visit, remembering to change sensors or charge devices, syncing multiple devices/applications, or doing routine checks/calibrations on devices)* |
| 2 | How much of a problem is the **annoyance and discomfort of your digital devices?** *(For example: device alarms or notifications interrupting daily activities or difficulty in wearing device with formal attire or beachwear)* |
| 3 | How much of a problem is the **time and effort required to solve problems with your digital devices?** *(For example: troubleshooting a new or old device, or finding educational resources from the healthcare team or device manufacturer)* |
| 4 | How much of a problem is it to take the **necessary precautions when using your digital devices?** *(For example: protecting your digital devices from drops, certain temperatures, or radiation (security checkpoint, MRIs, etc.) or carrying backups like a glucometer for continuous glucose monitors, or insulin pens for insulin pumps, etc.)* |
| 5 | How much of a problem is it **to give control of your health to digital devices?** *(For example: feeling uneasy or worried about the automatic functions of your device)* |
| 6 | 'The need for digital devices (including their visibility) on a regular basis **reminds me of my health problems**'. |

## DISCUSSION

We adapted the TBQ to capture the workload of using digital medicine tools and its negative impact on quality of life. The resulting instrument, TBQ+D, is a brief, self-reported measure of the overall burden of treatment inclusive of accessing and using digital medicine tools. Our findings demonstrate that digital burden is a distinct and meaningful component of treatment workload. By incorporating patient input throughout development, this work not only introduces a new tool to quantify digital burden in future research but also offers a patient-informed model for adapting existing measures to reflect modern care contexts.

### Comparison with Prior Research

Existing research has highlighted the potential for digital medicine tools to add to patients’ workload, particularly when these tools are poorly integrated or difficult to use.^17,34–37^ Our previously published work confirmed that digital self-management technologies can introduce unique forms of digital treatment burden, including device malfunctions, alert fatigue, data synchronization issues, and emotional distress.^26^

Despite growing recognition of this burden, no existing patient-reported instruments systematically capture the digital-specific components of treatment burden for patients with diabetes or one or more chronic conditions. Measures like the TBQ and PETS offer valuable insights into treatment burden but do not explicitly capture digital burden. This study addresses that gap by adapting the TBQ to include the burden of using digital tools directly informed by patient experiences.

### Implications for Clinical Practice

The development of the TBQ+D has two important implications for clinical practice. First, it offers a patient-informed instrument specifically designed to capture digital treatment burden among people with diabetes and, possibly, other chronic conditions— an increasingly important issue as chronic disease management becomes more reliant on digital tools. While additional validation is needed, the TBQ+D may help clinicians and researchers identify areas where digital tools contribute to care complexity, particularly for patients with high treatment demands.

Second, once validated, the TBQ+D can assist clinicians in recognizing that increased use of digital tools, even those meant to automate tasks, doesn’t always lead to a reduction in burden. Patients with more complex treatment needs may face a higher digital workload, highlighting the importance of a patient-centered approach to their implementation in practice.

### Limitations and Future Research

This study has two key limitations. First, it was conducted at a single academic medical center with a relatively homogenous patient population, which may limit the generalizability of the findings. Second, while the adaptation process included rigorous qualitative methods and patient input, this work focused on item development and cognitive testing.

The third phase in the process of developing TBQ+D involves its field testing, to test hypotheses that test is validity, i.e., its ability to truly capture the workload and negative effect on quality of life it may have induced by the work of accessing and using healthcare and enacting self-care tasks supported by digital tools. This process should continue with a range of other populations with chronic conditions and variable digital care use, thus advancing the evidence of the validity and responsiveness of the TBQ+D.

## CONCLUSION

This study describes the development of the Treatment Burden Questionnaire Plus Digital (TBQ+D), an adaptation of the original TBQ designed to capture burden associated with digital medicine tools. Grounded in patient experiences and refined through cognitive testing, the TBQ+D represents a step toward measuring burden of digital care in patients with living with diabetes.

By incorporating patient-centered design and directly addressing gaps in existing measures, the TBQ+D can support future research and clinical efforts to ensure digital tools reduce, rather than add to, the workload of living with diabetes. As the use of digital tools in care continues to grow, measuring burden of digital care will be essential to designing tools and workflows that promote patient centered care that is minimally disruptive.

## Data Availability

All data produced in the present work are contained in the manuscript and any additional data are available upon reasonable request to the authors.

## ACKNOWLEDGEMENTS

This work was funded by a competitive grant awarded by the Center for Digital Health at Mayo Clinic underwritten by the Noaber Foundation. The authors bear full responsibility for the content and any errors or omissions, and the views expressed do not necessarily reflect those of these entities.

The TBQ+D© is protected by international copyright, with all rights reserved to VMM and V-TT. For information on, or permission to use, the TBQ+D, please contact the authors.

## MEANING OF ACRYONMOMS

- **CGM:** Continuous Glucose Monitoring
- **EHR:** Electronic Health Record
- **IQR:** Interquartile Range
- **REDCap:** Research Electronic Data Capture
- **TBQ:** Treatment Burden Questionnaire

